# Associations of Combined Phenotypic Aging and Genetic Risk with Incident Cancer: A Prospective Cohort Study

**DOI:** 10.1101/2023.09.27.23296204

**Authors:** Lijun Bian, Zhimin Ma, Xiangjin Fu, Chen Ji, Tianpei Wang, Caiwang Yan, Juncheng Dai, Hongxia Ma, Zhibin Hu, Hongbing Shen, Lu Wang, Meng Zhu, Guangfu Jin

## Abstract

**Background:** Age is the most important risk factor for cancer, but aging rates are heterogeneous across individuals. We explored a new measure of aging-Phenotypic Age (PhenoAge)-in the risk prediction of site-specific and overall cancer.

**Methods:** Using Cox regression models, we examined the association of Phenotypic Age Acceleration (PhenoAgeAccel) with cancer incidence by genetic risk group among 374,463 participants from the UK Biobank. We generated PhenoAge using chronological age and 9 biomarkers, PhenoAgeAccel after subtracting the effect of chronological age by regression residual, and an incidence weighted overall cancer polygenic risk score (CPRS) based on 20 cancer site-specific polygenic risk scores (PRSs).

**Results:** Compared with biologically younger participants, those older had a significantly higher risk of overall cancer, with hazard ratios (HRs) of 1.22 (95% confidence interval, 1.18-1.27) in men, 1.26 (1.22-1.31) in women, respectively. A joint effect of genetic risk and PhenoAgeAccel was observed on overall cancer risk, with HRs of 2.29 (2.10-2.51) for men and 1.94 (1.78-2.11) for women with high genetic risk and older PhenoAge compared with those with low genetic risk and younger PhenoAge. PhenoAgeAccel was negatively associated with the number of healthy lifestyle factors (Beta = -1.01 in men, *P* < 0.001; Beta = -0.98 in women, *P* < 0.001).

**Conclusions:** Within and across genetic risk groups, older PhenoAge was consistently related to an increased risk of incident cancer with adjustment for chronological age and the aging process could be retarded by adherence to a healthy lifestyle.

## Introduction

Cancer continues to be the leading cause of death globally and the reduction of cancer-related deaths remains to be a public health priority (Bray et al., 2018). The morbidity and mortality of cancer increase dramatically with age, which demonstrated that aging is the greatest risk factor for cancer (Siegel, Miller, & Jemal, 2018). Although everyone gets older, individuals are aging at different rates (Rutledge, Oh, & Wyss-Coray, 2022). Therefore, the variation in the pace of aging between-person may reflect the differences in susceptibility to cancer and death. Thus, measurement of an individual’s biological age, particularly at the early stage of life, may promote the primary and secondary prevention of cancer through earlier identification of high risk groups.

Recently, Morgan and colleagues developed and validated a novel multi-system-based aging measurement (Levine et al., 2018), Phenotypic Age (PhenoAge), which has been shown to capture long-term vulnerability to diseases like COVID-19, and strongly predict morbidity and mortality risk in diverse populations (Kuo, Pilling, Atkins, et al., 2021; Liu et al., 2018). However, it is largely unknown whether PhenoAge can predict overall cancer risk and identify high risk individuals for potential personalized prevention.

To date, more than 2,000 genetic loci have been identified as susceptibility markers for certain cancers by genome-wide association studies (GWAS) (Buniello et al., 2019). Although the effect of these individual loci is relatively modest on cancer risk, a polygenic risk score (PRS) combining multiple loci together as an indicator of genetic risk has been proved to effectively predict incidence of site-specific cancer (Dai et al., 2019; Lecarpentier et al., 2017; Mars et al., 2020). Recently, we systematically created site-specific cancer PRS for 20 cancer types, and constructed an incidence-weighted cancer polygenic risk score (CPRS) to assess the effect of genetic risk on overall incident cancer risk based on the UK Biobank (Zhu et al., 2021). Previous study had indicated an interaction between genetic factor and age on cancer risk (Mavaddat et al., 2015). However, the extent to interaction between genetic factor and PhenoAge on overall cancer risk remained unclear.

In this study, we calculated PhenoAge in accordance with the method described previously and then evaluated the effectiveness of PhenoAge in predicting risk of overall cancer in the UK Biobank. We also assessed the extent to which a level of accelerated aging was associated with an increased overall cancer risk across groups with a different genetic risk defined by the CPRS.

## Methods

### Participants

The participants included in this study are derived from the UK Biobank. The UK Biobank is a large population-based cohort study and detail protocol is publicly available (Bycroft et al., 2018). In brief, approximately 500,000 participants aged 40-70 were recruited from 22 assessment centers across England, Scotland, and Wales between 2006 and 2010 at baseline. Each eligible participant completed a written informed consent form and provided information on lifestyle and other potentially health-related aspects through extensive baseline questionnaires, interviews, and physical measurements. Meanwhile, biological samples of participants were also collected for biomarker assays and a blood draw was collected for genotyping. The UK Biobank study has approval from the Multi-center Research Ethics Committee, the National Information Governance Board for Health and Social Care in England and Wales, and the Community Health Index Advisory Group in Scotland (http://www.ukbiobank.ac.uk/ethics/).

### PhenoAge and PhenoAgeAccel PRS

We calculated PhenoAge in accordance with the method described previously (Levine et al., 2018). Briefly, PhenoAge was calculated based on mortality scores from the Gompertz proportional hazard model on chronological age and 9 multi-system clinical chemistry biomarkers (albumin, creatinine, glucose, [log] C-reactive protein [CRP], lymphocyte percent, mean cell volume, red blood cell distribution width, alkaline phosphatase, and white blood cell count) to predict all-cause mortality. The Biomarkers in the UK Biobank were measured at baseline (2006-2010) for all participants. To correct distribution skewness, we set the top and bottom 1% of values to the 99th and 1st percentiles. The formula of PhenoAge is given by

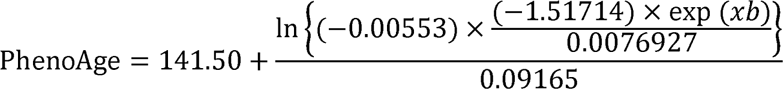

where

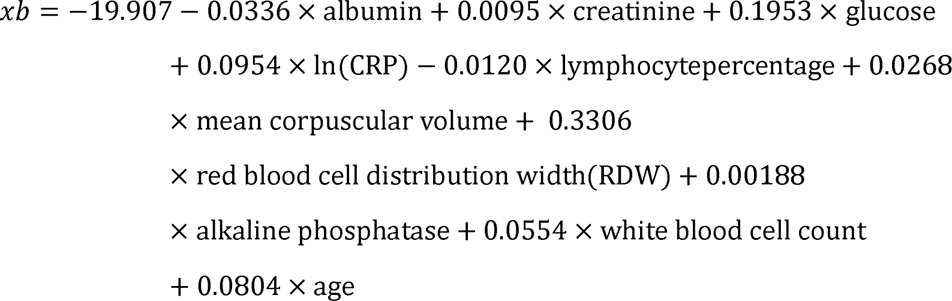

Finally, we calculated Phenotypic Age Acceleration (PhenoAgeAccel), which was defined as the residual resulting from a linear model when regressing Phenotypic Age on chronological age. Therefore, PhenoAgeAccel represents Phenotypic Age after accounting for chronological age (i.e., whether a person appears older [positive value] or younger [negative value] than expected, biologically, based on his/her age).

55 independent PhenoAgeAccel-associated SNPs (*P* < 5 × 10^-8^) and corresponding effect sizes were derived from a large-scale PhenoAgeAccel GWAS including 107,460 individuals of European ancestry (Kuo, Pilling, Liu, Atkins, & Levine, 2021). A PhenoAgeAccel PRS was created using an additive model as previously described (Dai et al., 2019). In short, the genotype dosage of each risk allele for each individual was summed after multiplying by its respective effect size of PhenoAgeAccel.

### PRS calculation and CPRS construction

The calculation of site-specific cancer PRSs and the construction of CPRSs have been described in our previous published study (Zhu et al., 2021). In brief, for individual cancer, risk associated single nucleotide polymorphisms (SNPs) and corresponding effect sizes were derived from the largest published GWASs in terms of sample size. Next, the dosage of each risk allele for each individual was summed after multiplication with its respective effect size of site-specific cancer. Except for non-melanoma skin cancer and those without relevant GWAS or significant genetic loci published by now, we derived PRSs for 20 cancer types in this analysis. To generate an indicator of genetic risk for overall cancer, we constructed the CPRS as follows:

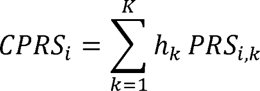

Where *CPRS_i_* is the cancer polygenic risk score of i^th^ individual, *h_k_* is the age-standardized incidence of site-specific cancer *k* in UK population, and *PRS_i,k_* is the aforementioned PRS of site-specific cancer *k*. Given the different spectrum of cancer incidence between men and women, CPRS were constructed for males and females, respectively.

### Assessment of healthy lifestyle

We adopted five healthy lifestyle factors according to the World Cancer Research Fund/American Institute of Cancer Research recommendations (https://www.aicr.org/cancer-prevention/) (Shams-White et al., 2019), i.e., no current smoking, no alcohol consumption, regular physical activity, moderate BMI (body-mass index, 18.5∼30), and a healthy diet pattern. Participants of no current smoking were defined as never smoker or former smokers who had quit smoking at least 30 years. No alcohol consumption was defined as never alcohol use. Regular physical activity was defined as at least 75 minutes of vigorous activity per week or 150 minutes of moderate activity per week (or an equivalent combination) or engaging in vigorous activity once and moderate physical activity at least 5 days a week (Lourida et al., 2019). A healthy diet pattern was ascertained consumption of an increased amount of fruits, vegetables, whole grains, fish and a reduced amount of red meats and processed meats (Lourida et al., 2019). The lifestyle index ranged from 0 to 5, with higher index indicating a healthier lifestyle.

### Outcomes

Outcomes of incident cancer events in the UK Biobank were ascertained through record electronic linkage with the National Health Service central registers and death registries in England, Wales and Scotland. Complete follow-up was updated to 31 October 2015 for Scotland, and to 31 March 2016 for England & Wales. Cancer events were coded using the 10^th^ Revision of the International Classification of Diseases. The outcome of all cancer events were obtained from data field 40006 and 40005 of the UK Biobank.

### Statistical analysis

Cancer risk of participants in the UK Biobank was assessed from baseline until to the date of diagnosis, death, loss to follow-up, or date of complete follow-up, whichever occurred first. Multivariable Cox proportional hazards regression analyses were performed to assess associations between PhenoAgeAccel and cancer risk and to estimate hazard ratios (HRs) as well as 95% confidence intervals (CI). Schoenfeld residuals and log-log inspection were used to test the assumption of proportional hazards. HRs associated with per 5 years increased of PhenoAgeAccel was calculated for site-specific cancer and overall cancer respectively. We compared HRs between biologically younger and older participants. In addition, we calculated HRs for participants at low (the bottom quintile of PhenoAgeAccel), intermediate (quintiles 2-4), and high (the top quintile) accelerated aging, and HRs for participants splitted by decile of accelerated aging.

Meanwhile, participants were also divided into low (the bottom quintile of CPRS), intermediate (quintiles 2-4), and high (the top quintile) genetic risk groups. Absolute risk within each subgroup defined by PhenoAgeAccel and CPRS were calculated as the percentage of incident cancer cases occurring in a given group. We calculated absolute risk increase as the difference in cancer incidences among given groups, extrapolated the difference in 5-year event rates among given groups. The 95% CIs for the absolute risk increase were derived by drawing 1,000 bootstrap samples from the estimation dataset. We performed additive interaction analysis between genetic risk (defined by CPRS) and PhenoAgeAccel on overall cancer risk, as well as genetic risk (defined by PhenoAgeAccel PRS) and lifestyle on PhenoAgeAccel using two indexes: the relative excess risk due to interaction (RERI) and the attributable proportion due to interaction (AP) (R. Li & Chambless, 2007). The 95% CIs of the RERI and AP were estimated by bootstrap (N = 5,000), which would contain 0 if there was no additive interaction. We also used multivariable linear regression models to assess associations between the PhenoAgeAccel and individual lifestyle factors with adjustment for age, family history of cancer, Townsend deprivation index, height, and the first 10 principal components of ancestry. All the above mentioned analyses were performed for men and women separately.

Participants with missing data on any of the covariates were multiple imputed, and independent analyses were also performed based on complete data for sensitivity analyses. Besides, to examine the reliability of our results, we conducted several sensitivity analyses: (1) reclassifying PhenoAgeAccel levels based on quartiles (bottom, 2-3, and top quartiles defined as low, intermediate, and high accelerated aging, respectively) or tertiles (corresponding to low, intermediate, and high accelerated aging) of PhenoAgeAccel; (2) reevaluating the effect of PhenoAgeAccel based on participants of unrelated British ancestry; (3) excluding incident cases of any cancer occurring during the two years of follow-up; and (4) retrained PhenoAge in cancer-free participants based on mortality. All *P* values were two-sided and *P* < 0.05 was considered statistically significant. All statistical analyses were performed with R software, version 3.6.3 (R Project for Statistical Computing).

## Results

### Participants

After removing participants who had withdrawn their consent, had been diagnosed with cancer before baseline, failed to be genotyped, reported a mismatch sex with genetic data, or with missing data on PhenoAge, the final analytic dataset included 374,463 eligible participants (173,431 men and 201,032 women). Of which, 169,573 participants were biologically older, with 92,189 men and 77,384 women, whose median PhenoAgeAccel were 3.28 (interquartile range [IQR]: 1.50 to 6.06) and 3.07 (IQR: 1.33 to 5.79) respectively; 204,890 participants were biologically younger, with 81,242 men and 123,648 women, whose median PhenoAgeAccel were -2.61 (IQR: -4.35 to -1.25) and -3.55 (IQR: -5.64 to -1.81) respectively (**Table 1**, **Appendix 1-figure 1).**

**Table 1.** Baseline characteristics of participants stratified by PhenoAgeAccel categories.

|  | Men (n= 173431) |  | Women (n= 201032) |  |
| --- | --- | --- | --- | --- |
|  | Biologically younger<br>n=81242 | Biologically older<br>n=92189 | Biologically younger<br>n=123648 | Biologically older<br>n=77384 |
| **PhenoAgeAccel, median (IQR), years** | -2.61 (-4.35--1.25) | 3.28 (1.50-6.06) | -3.55 (-5.64--1.81) | 3.07 (1.33-5.79) |
| **Age at baseline, median (IQR), years** | 57.00 (49.00-62.00) | 58.00 (50.00-64.00) | 58.00 (51.00-63.00) | 56.00 (48.00-62.00) |
| **Height, median (IQR), centimeters** | 176.00 (171.00-181.00) | 175.00 (171.00-180.00) | 162.50 (158.00-167.00) | 162.00 (158.00-166.50) |
| **Townsend deprivation index, median (IQR)** | -2.34 (-3.76-0.16) | -1.86 (-3.52-1.10) | -2.33 (-3.73-0.07) | -1.79 (-3.44-1.10) |
| **Family history of cancer, n (%)** |  |  |  |  |
| No | 53907 (66.35) | 60654 (65.79) | 79800 (64.54) | 50924 (65.81) |
| Yes | 27335 (33.65) | 31535 (34.21) | 43848 (35.46) | 26460 (34.19) |
| **Healthy lifestyle factors, n (%)** |  |  |  |  |
| No current smoking | 46412 (57.13) | 43216 (46.88) | 78743 (63.68) | 45416 (58.69) |
| No alcohol consumption | 2064 (2.54) | 2828 (3.07) | 6221 (5.03) | 5459 (7.05) |
| Normal BMI | 66120 (81.39) | 62654 (67.96) | 103054 (83.34) | 49003 (63.32) |
| Regular physical activity | 54346 (66.89) | 57733 (62.62) | 80629 (65.21) | 46468 (60.05) |
| Healthy diet | 18540 (22.82) | 15486 (16.80) | 39547 (31.98) | 19826 (25.62) |
| **Healthy lifestyle<sup>a</sup>, n (%)** |  |  |  |  |
| Favorable | 7781 (9.58) | 5255 (5.70) | 17781 (14.38) | 7178 (9.28) |
| Intermediate | 57683 (71.00) | 57489 (62.36) | 87272 (70.58) | 49324 (63.74) |
| Unfavorable | 15778 (19.42) | 29445 (31.94) | 18595 (15.04) | 20882 (26.98) |
| **Albumin, median (IQR), (g/L)** | 46.20 (44.62-47.85) | 44.95 (43.28-46.64) | 45.47 (43.87-47.12) | 44.04 (42.42-45.70) |
| **Alkaline phosphatase, median (IQR), (U/L)** | 75.00 (64.30-87.40) | 82.80 (70.10-97.90) | 78.70 (65.10-93.90) | 86.20 (70.60-104.00) |
| **Creatinine, median (IQR), (umol/L)** | 77.60 (70.90-84.70) | 82.50 (74.40-91.60) | 61.70 (56.00-67.90) | 65.50 (58.90-73.20) |
| **Glucose, median (IQR), (mmol/l)** | 4.82 (4.49-5.14) | 5.09 (4.72-5.60) | 4.84 (4.54-5.14) | 5.05 (4.70-5.55) |
| **C-reactive protein, median (IQR), (mg/dL)** | 0.09 (0.05-0.16) | 0.19 (0.10-0.36) | 0.10 (0.05-0.20) | 0.25 (0.12-0.51) |
| **Lymphocyte percent, median (IQR), (%)** | 29.86 (25.48-34.48) | 25.60 (21.22-30.24) | 31.09 (26.58-35.80) | 27.00 (22.62-31.61) |
| **Mean cell volume, median (IQR), (fL)** | 90.90 (88.46-93.30) | 91.90 (89.10-94.74) | 91.10 (88.60-93.54) | 90.98 (87.64-94.02) |
| **Red cell distribution width, median (IQR), (%)** | 13.01 (12.70-13.37) | 13.63 (13.21-14.11) | 13.10 (12.71-13.50) | 13.90 (13.40-14.55) |
| **White blood cell count, median (IQR), (1000 cells/uL)** | 6.12 (5.29-7.10) | 7.27 (6.17-8.51) | 6.24 (5.37-7.26) | 7.41 (6.30-8.73) |

<sup>a</sup> Healthy lifestyle was defined as favorable (4-5 healthy lifestyle factors), intermediate (2-3 healthy lifestyle factors), and unfavorable (0-1 healthy lifestyle factor).

### Associations of PhenoAgeAccel with cancer risk

There were 22,370 incident cancer cases, with 11,532 men and 10,838 women, during a median follow-up of 7.09 years (IQR: 6.35 to 7.72). The PhenoAgeAccel was significantly associated with increased risk for cancer sites of lip-oral cavity-pharynx, esophagus, stomach, colon-rectum, pancreas, lung, breast, cervix uteri, corpus uteri, prostate, kidney, bladder, multiple myeloma, Hodgkin’s disease, and lymphoid leukaemia, while negatively associated with risk of prostate cancer after adjusting for chronological age and other covariates (**Figure 1**, **Appendix 1-table 1**).

**Figure 1.**
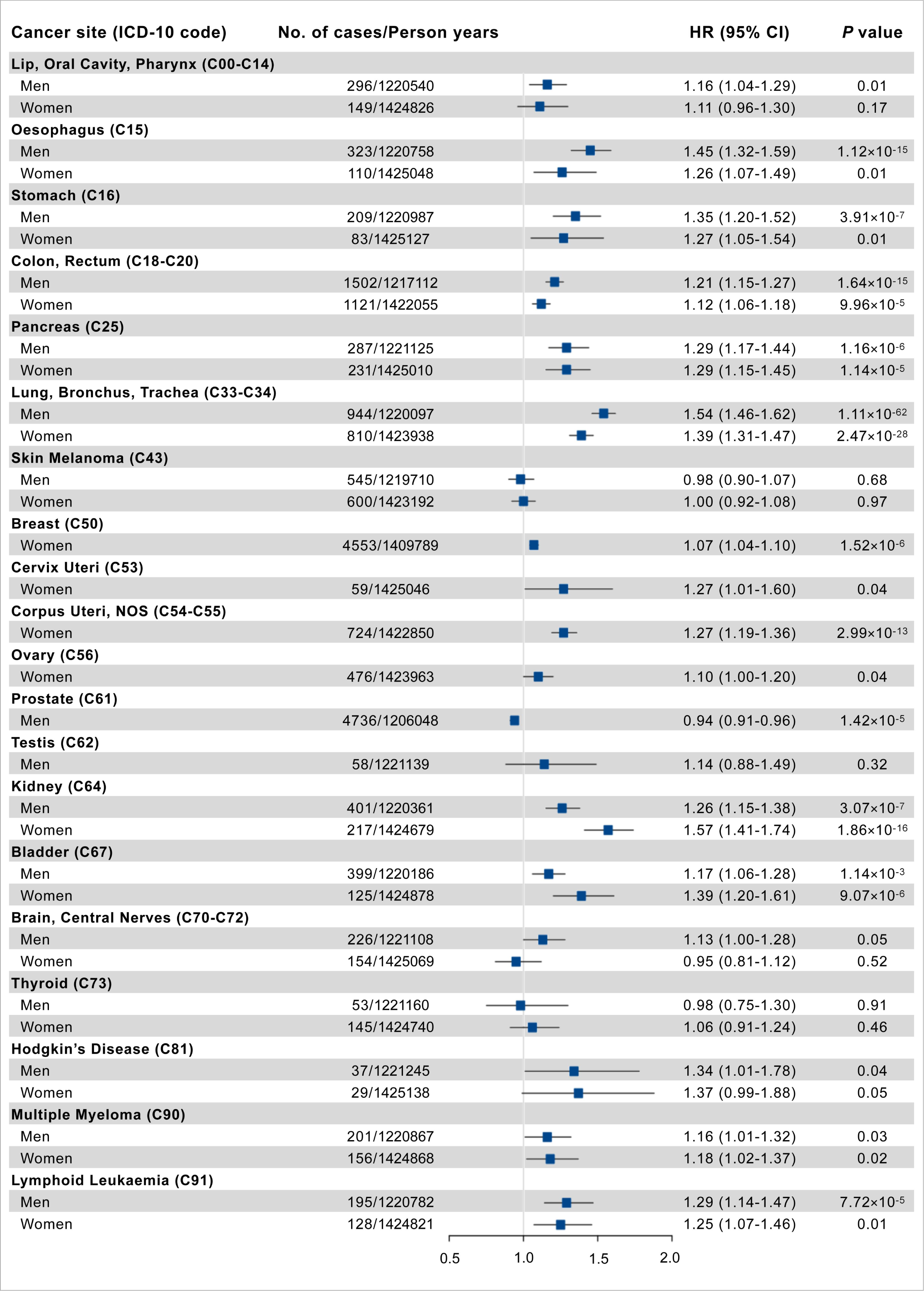
Association results of PhenoAgeAccel with site-specific cancer risk per 5 years increased. Cox proportional hazards regression adjusted for age, height, cancer family history, Townsend deprivation index at recruitment, and the first 10 principal components of ancestry.

For overall cancer, we observed an obviously higher distribution of PhenoAgeAccel in incident cancer cases than participants without incident cancer in both men and women (**Figure 2A** and **B**). PhenoAgeAccel was significantly associated with an increased risk of overall cancer in men (HR = 1.15, 95% CI, 1.13-1.17, *P* < 0.0001) and women (HR = 1.15, 95% CI, 1.13-1.17, *P* < 0.0001) per 5 years increase (**Table 2**). We also observed a significantly gradient increase in incident cancer risk from decile 1 to decile 10 of PhenoAgeAccel (**Figure 2C** and **D**). Compared with biologically younger participants, those older had a significantly higher risk of overall cancer, with HRs of 1.22 (95% CI, 1.18-1.27, *P* < 0.0001) in men, 1.26 (95% CI, 1.22-1.31, *P* < 0.0001) in women, respectively (**Figure 2E** and **F**). Besides, Compared with individuals at low accelerated aging (the bottom quintile of PhenoAgeAccel), those in the intermediate (quintiles 2 to 4) and high accelerated aging (the top quintile) had a significantly higher risk of overall cancer, with HRs of 1.15 (95% CI, 1.09-1.21, *P* < 0.0001) and 1.44 (95% CI, 1.36-1.53, *P* < 0.0001) in men, 1.15 (95% CI, 1.09-1.21, *P* < 0.0001) and 1.46 (95% CI, 1.38-1.55, *P* < 0.0001) in women, respectively. These results did not change after adjustment for genetic risk and lifestyle factors (**Table 2**). Similar patterns were noted in a series of sensitivity analyses with reclassifying accelerated aging levels according to quartiles or tertiles of the PhenoAgeAccel (**Appendix 1-table 2**), exclusion of incident cancer cases occurred during the two years of follow-up (**Appendix 1-table 3**), in the unimputed data (**Appendix 1-table 4**), in the unrelated British population (**Appendix 1-table 5**), or using retrained PhenoAge in cancer-free participants (**Appendix 1-table 6**).

**Figure 2.**
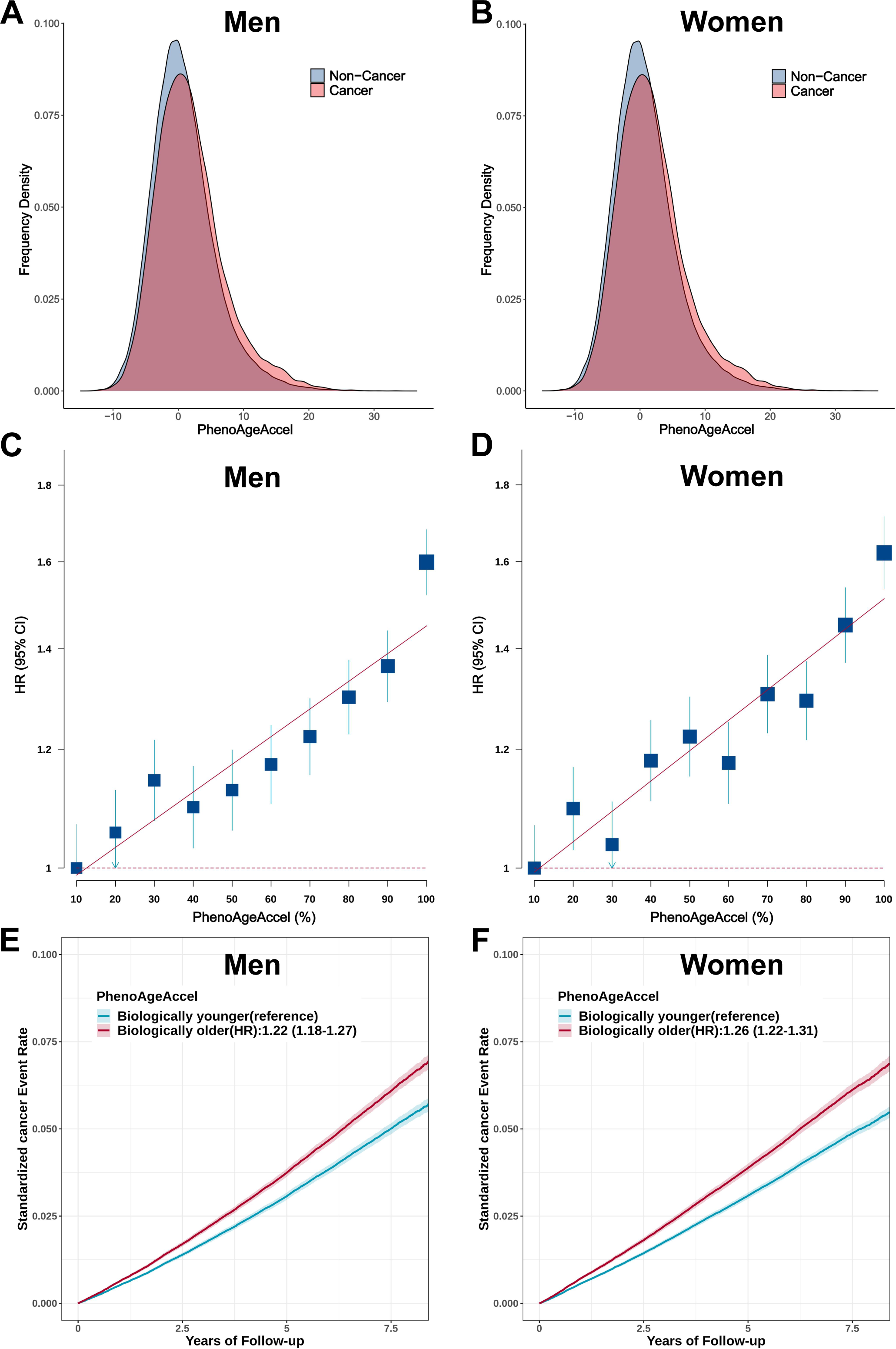
Effect of PhenoAgeAccel on the risk of incident cancer in the UK Biobank. The distrubution of PhenoAgeAccel between participants with incident cancer and those without incident cancer in the UK Biobank for men(A) and women (B). Participants in the UK Biobank were divided into ten equal groups according to the PhenoAgeAccel for men (C) and women (D), and the hazard ratios (HRs) of each group were compared with those in the bottom decile of PhenoAgeAccel. Error bars are 95% CIs. Standardized rates of cancer events in younger and older PhenoAge groups in the UK Biobank for men (E) and women (F). HRs and 95% CIs were estimated using Cox proportional hazard models with adjustment for age, height, family history of cancer, Townsend deprivation index, and the first 10 principal components of ancestry. Shaded areas are 95% CIs.

**Table 2.**
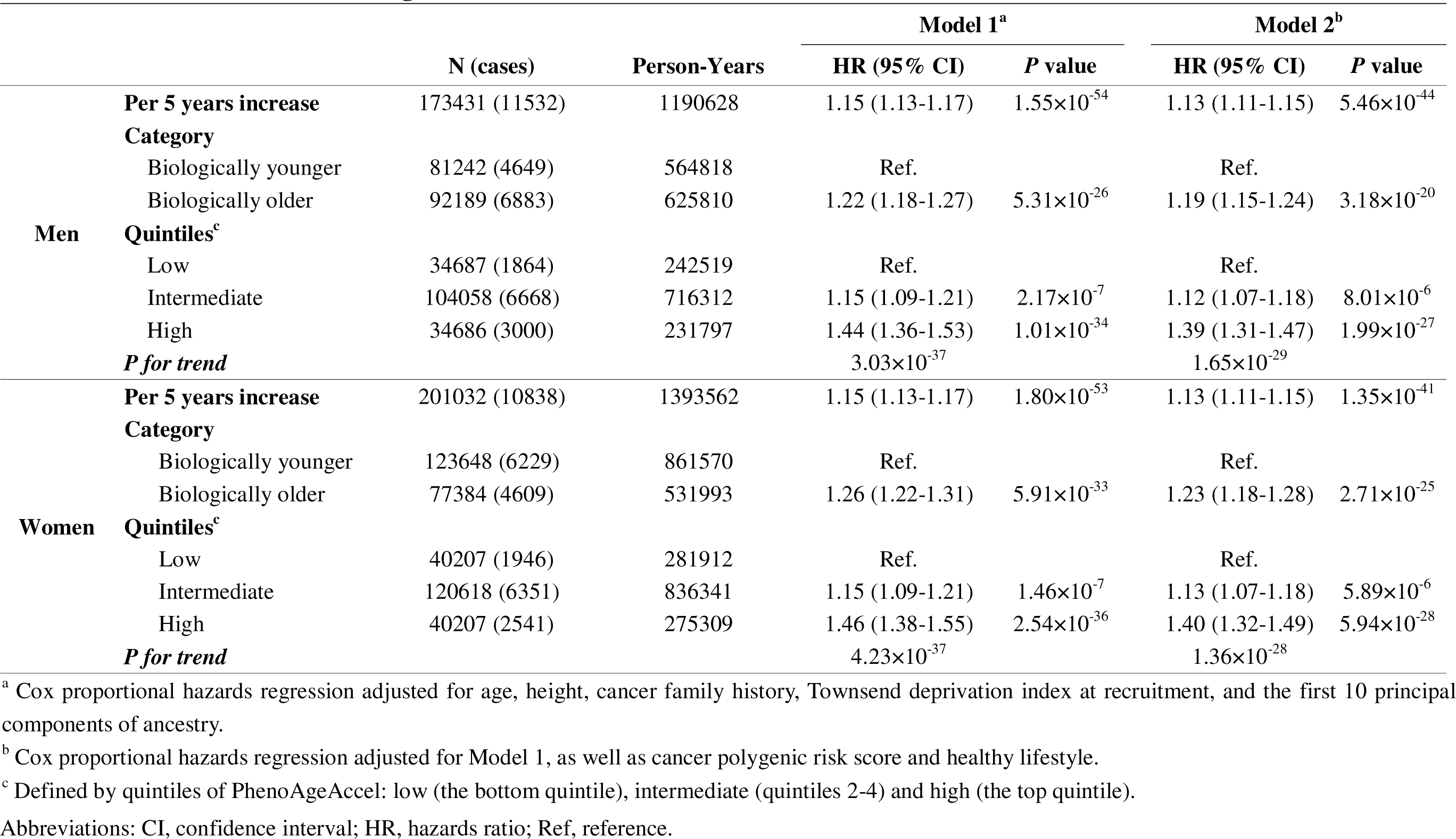
Association between P henoAgeAccel and cancer risk.

### Joint effect and interaction of genetic factor and PhenoAgeAccel on overall cancer risk

The overall incident cancer risk associated with both genetic risk and PhenoAgeAccel in a dose-response manner (**Figure 3**). Of participants with high genetic risk and older PhenoAge, the incidence rates of overall cancer per 100,000 person-years were estimated to be 1477.89 (95% CI, 1410.87-1544.92) in men and 1076.17 (95% CI, 1014.14-1138.19) in women versus 581.06 (95% CI, 537.12-625.00) in men and 594.71 (95% CI, 558.50-630.92) in women with low genetic risk and younger PhenoAge. Approximate double risks [HR, 2.29 (95% CI, 2.10-2.51) in men, *P* < 0.0001; 1.94 (95% CI, 1.78-2.11) in women, *P* < 0.0001] were observed in participants with high genetic risk and older PhenoAge, compared with those with low genetic risk and younger PhenoAge. Similar patterns were noted by reclassifying accelerated aging levels into low (the bottom quintile of PhenoAgeAccel), intermediate (quintiles 2-4), and high (the top quintile) (**Appendix 1-figure 2**). However, we did not observe interaction between genetic and PhenoAgeAccel on overall cancer risk in men and women (**Appendix 1-table 7**).

**Figure 3.**
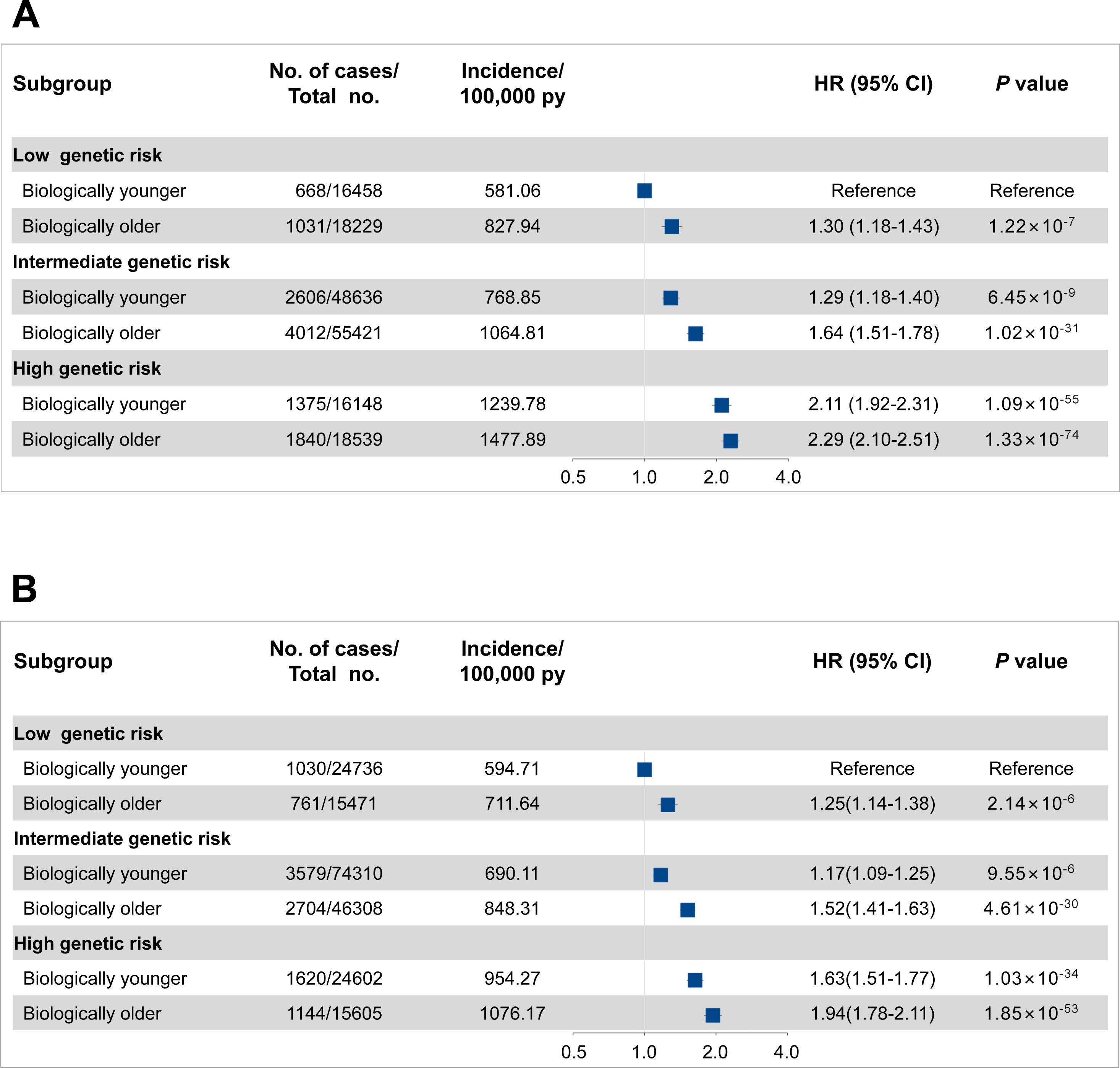
Risk of incident cancer according to genetic and PhenoAgeAccel categories in the UK Biobank for men (A) and women (B). The HRs were estimated using Cox proportional hazard models with adjustment for age, height, family history of cancer, Townsend deprivation index, and the first 10 principal components of ancestry. Participants were divided into younger and older PhenoAge under different genetic risk groups.

### Disadvantages of older PhenoAge with overall incident cancer

In further stratification analyses by genetic risk category with younger PhenoAge as the reference group, we confirmed that older PhenoAge was significantly associated with a higher incident cancer risk across genetic risk groups (**Table 3**). Among participants at high genetic risk, the standardized 5-year incident cancer rates were 5.78% and 4.58% for biologically younger men and women versus 6.90% and 5.17% for those older, respectively. Similarly, among participants at low genetic risk, the standardized 5-year incident cancer rates increased from 2.71% and 2.83% for biologically younger to 3.87% and 3.39% for those older in men and women, respectively. Similar patterns were noted by reclassifying accelerated aging levels into low (the bottom quintile of PhenoAgeAccel), intermediate (quintiles 2-4), and high (the top quintile) (**Appendix 1-table 8**).

**Table 3.** Risk of incident cancer according to PhenoAgeAccel categories within each genetic risk level^a^.

| Gender | PhenoAgeAccel category | Low genetic risk |  | Intermediate genetic risk |  | High genetic risk |  |
| --- | --- | --- | --- | --- | --- | --- | --- |
|  |  | Biologically younger | Biologically older | Biologically younger | Biologically older | Biologically younger | Biologically older |
| Men | No. of cases/Person years | 668/114962 | 1031/124526 | 2606/338950 | 4012/376782 | 1375/110907 | 1840/124502 |
|  | Hazards ratio (95% CI) | Ref. | 1.29 (1.17-1.42) | Ref. | 1.27 (1.21-1.33) | Ref. | 1.10 (1.02-1.18) |
|  | *P* value |  | 3.53×10<sup>-7</sup> |  | 3.15×10<sup>-21</sup> |  | 1.07×10<sup>-2</sup> |
|  | Absolute risk (%) - 5 years (95% CI) | 2.71 (2.49-2.94) | 3.87 (3.60-4.14) | 3.57 (3.41-3.72) | 4.95 (4.78-5.13) | 5.78 (5.44-6.11) | 6.90 (6.55-7.26) |
|  | Absolute risk increase (%) - 5 years (95% CI) | Ref. | 1.16 (0.84-1.46) | Ref. | 1.39 (1.19-1.60) | Ref. | 1.13 (0.68-1.52) |
| Women | No. of cases/Person years | 1030/173195 | 761/106937 | 3579/518611 | 2704/318753 | 1620/169764 | 1144/106303 |
|  | Hazards ratio (95% CI) | Ref. | 1.25 (1.14-1.38) | Ref. | 1.30 (1.24-1.37) | Ref. | 1.19 (1.10-1.28) |
|  | *P* value |  | 2.65×10<sup>-6</sup> |  | 1.84×10<sup>-24</sup> |  | 8.19×10<sup>-6</sup> |
|  | Absolute risk (%) - 5 years (95% CI) | 2.83 (2.63-3.02) | 3.39 (3.12-3.65) | 3.27 (3.15-3.39) | 4.02 (3.86-4.19) | 4.58 (4.33-4.83) | 5.17 (4.84-5.49) |
|  | Absolute risk increase (%) - 5 years (95% CI) | Ref. | 0.56 (0.26-0.86) | Ref. | 0.76 (0.57-0.93) | Ref. | 0.59 (0.23-0.98) |

<sup>a</sup> Cox proportional hazards regression is adjusted for age, height, family history of cancer, Townsend deprivation index, height and the first 10 principal components of ancestry.
Abbreviations: CI, confidence interval; Ref, reference.

In addition, to evaluate the implication for cancer screening in populations with different PhenoAgeAccel, we estimated the 5-year absolute risk of overall cancer between biologically younger and older participants with the increasing of age. Assuming 2% of absolute risk within the next 5 years as the threshold to be recommended for cancer screening, biologically younger men would reach the threshold at age 52, whereas those older men would reach the threshold at age 50; similarly, biologically younger women would reach the 2% of 5-year absolute risk at age 46, whereas those older women would reach the threshold at age 44 (**Figure 4**).

**Figure 4.**
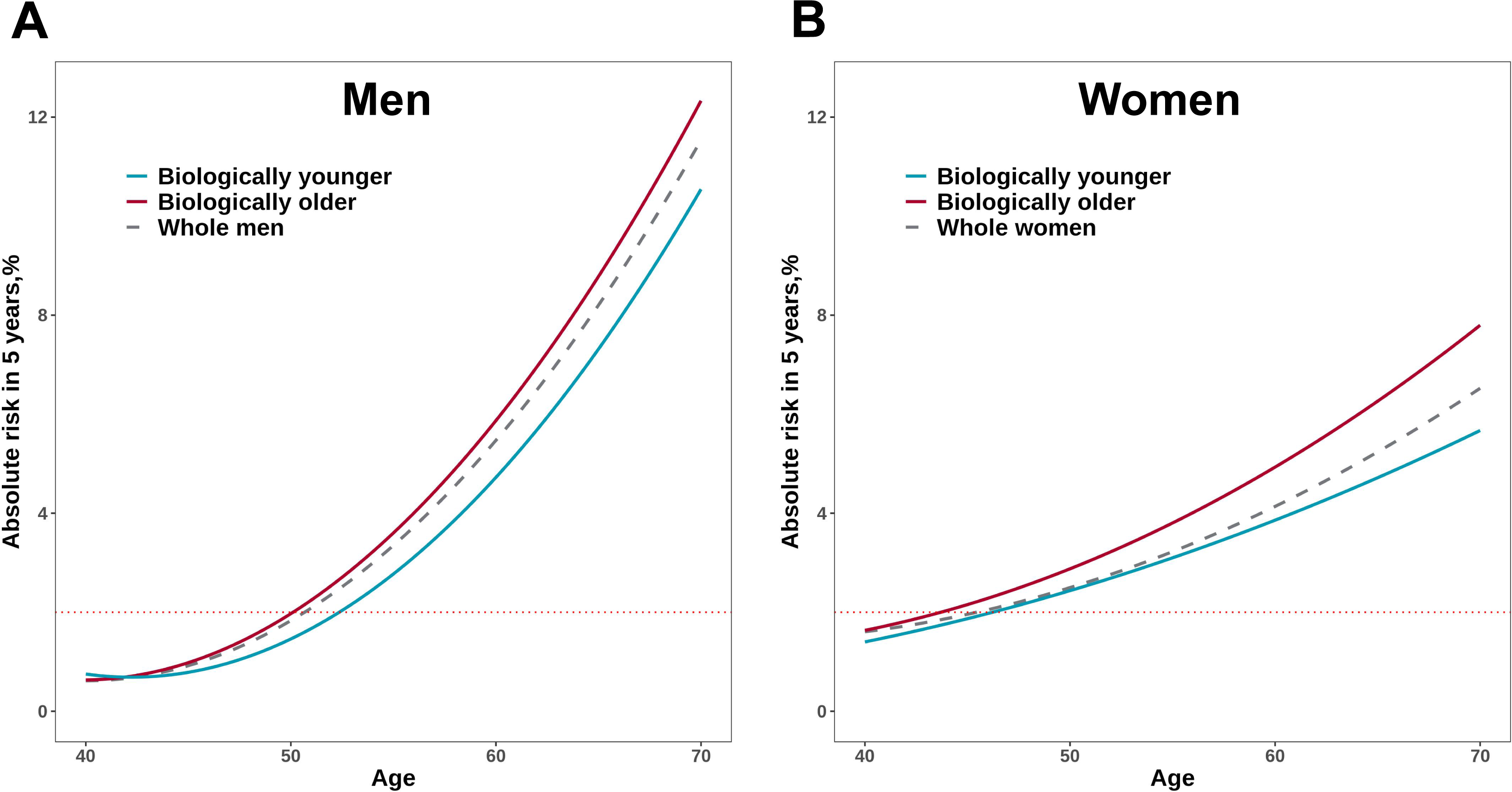
Absolute risk estimates of overall cancer based on the UK Biobank for men (A) and women (B). The x-axis is chronological age. The curves describe average risk of participants in younger and older PhenoAge groups. The dashed curve represents the average risk of the whole population in different ages. The red horizontal dotted line represents 2% of 5-year absolute risks of overall cancer.

### Associations of lifestyle factors with PhenoAgeAccel

In the UK Biobank, biologically younger men (9.6%, 7,781/81,242) and women (14.4%, 17,781/123,648) were more likely to have favorable lifestyle (4 to 5 healthy lifestyle factors) than older men (5.7%, 5,255/92,189) and women (9.3%, 7,178/77,384) (**Table 1**). Among both men and women, we observed that PhenoAgeAccel decreased with the increase of healthy lifestyle factors (**Appendix 1-table 9**). In addition, we found a significant negative correlation between the number of healthy lifestyle factors and PhenoAgeAccel (Beta = -1.01 in men, *P* < 0.001; Beta = -0.98 in women, *P* < 0.001) (**Appendix 1-table 10**). However, we did not observe any interaction between genetic risk and lifestyle on PhenoAgeAccel in both men and women (**Appendix 1-table 11**).

## Discussion

In this study, we calculated PhenoAgeAccel to explore the effect of accelerated aging on the risk of cancer, and demonstrated a positive association between accelerated aging and increased cancer risk after adjustment for chronological age in the UK Biobank. Meanwhile, older PhenoAge was consistently associated with an increased absolute risk of incident cancer within each genetic risk group; and participants with high genetic risk and older PhenoAge had the greatest incident cancer risk. Therefore, our findings provided the evidence for PhenoAgeAccel to be used for risk stratification of cancer, which were independent from genetic risk. Moreover, we also demonstrated that participants with older biological age often reaches the screening threshold 2 years in advance compared with biologically younger peers; and keeping a healthy lifestyle can effectively slow down the aging process.

Older age has been long recognized as the main risk factor for cancer, and the multistage model of carcinogenesis posits that the exponential increase in cancer incidence with age were mainly resulted from the sequential accumulation of oncogenic mutations in different tissues throughout life (Laconi, Marongiu, & DeGregori, 2020). In consistent with this, age and exposure (i.e. smoking, ultraviolet light) dependent mutation signatures have been identified in several cancers by tissue sequencing (Alexandrov et al., 2020). However, biological aging is an enormously complex process and is thought to be influenced by multiple genetic and environmental factors (van Dongen et al., 2016). Therefore, several biomarkers, i.e. ‘ageing clocks’ derived from epigenomic, transcriptomic, proteomic and metabolomic data, have been proposed to measure the biological age and predict risk of cancer and other diseases (Rutledge et al., 2022; Zhang et al., 2022). However, these measures were usually based on omics data and was not suitable for application in large population by now. As a result, results from this study would provide a cost-effective indicator for measuring of biological age as well as a novel biomarker for cancer risk prediction.

The associations between biological age and cancer risk has been investigated by several studies recently. Li et al. explored three DNA methylation phenotypic age and cancer risk in four subsets of a population-based cohort from Germany, and reported strong positive associations for lung cancer, while strong inverse associations for breast cancer (X. Li, Schottker, Holleczek, & Brenner, 2022). Meanwhile, results from Melbourne Collaborative Cohort Study reported that epigenetic aging was associated with increased cancer risk of kidney cancer and B-cell lymphoma (Dugue et al., 2018). However, because of sample size, the association results were still inconsistent for DNA methylation phenotypic age among different studies. Leukocyte telomere length was also significantly associated with age and were regarded as an indicator of aging. Based on data from the UK Biobank, Schneider et al. recently explored the associations between telomere length and risk of several disease, and reported significant positive associations of telomere length for lymphoid leukemia, multiple myeloma, non-Hodgkin lymphoma, esophagus cancer, while negative associations for malignant neoplasm of brain, mesothelioma, and melanoma (Schneider et al., 2022). The positive associations were in consistent with our findings, however, the negative associations were not significant in our study. Meanwhile, the study did not indicate associations for other cancers, including cancers of lung, stomach, pancreases, and kidney, which showed relatively large effects (HR > 1.3) in our study. These findings indicated that the different measures of biological age may reflect the different aspects of aging, and could be joint application in cancer risk assessment.

Recently, several studies have confirmed the associations between PhenoAgeAccel and cancer risk. Mak et al. explored three measures of biological age, including PhenoAge, and assessed their associations with the incidence of overall cancer and five common cancers (breast, prostate, lung, colorectal, and melanoma) (Mak et al., 2023). In our previous study, we investigated the association between PhenoAgeAccel and lung cancer risk and analyzed the joint and interactive effects of PhenoAgeAccel and genetic factors on the risk of lung cancer (Ma et al., 2023). In comparison to these studies, our analysis expanded the range of cancers to 20 types and further explored the associations in different genetic and lifestyle contexts. Moreover, we also evaluated the potential implications of PhenoAge in population-level cancer screening. In addition, we observed a negative association between PhenoAgeAccel and prostate cancer risk. The unexpected association may have been confounded by diabetes and altered glucose metabolism, both of which are closely linked to aging. When we removed HbA1c and serum glucose from the biological age algorithms, the association became non-statistically significant. Similar findings were also reported by Mak et al. (Mak et al., 2023) and Dugue et al. (Dugue et al., 2021).

The associations between PhenoAgeAccel and increased cancer risk may be partly attribute to a result of decline in the immune system and accumulation of environmental carcinogenic factors. Recent GWASs of PhenoAgeAccel showed that SNPs associated with PhenoAgeAccel were enriched in pathways of immune system and activation of pro-inflammatory (Kuo, Pilling, Liu, Atkins, & Levine, 2021; Levine et al., 2018). In addition to genetics, behaviors (i.e. obesity, smoking, alcohol consumption, and physical activity), and life course circumstances (i.e. socioenvironmental circumstances during childhood and adulthood) were reported to account for about 30% variances of phenotypic aging (Liu et al., 2019). This was in accordance with our findings that, adherence to healthy lifestyles (involving no current smoking, normal BMI, regular physical activity, and healthy diet) could slow down the aging process. In other words, these healthy lifestyles considered in our and previous studies may be causal drivers of phenotypic aging, they represent a more targetable strategy for reducing overall cancer burden by retarding the aging process. Therefore, PhenoAge provide a meaningful intermediate phenotype that can be used to guide interventions for high risk groups and track intervention efficacy (Liu et al., 2019).

This study has several strengths, including a large sample size, a prospective design of the UK Biobank study, and an effective application of PhenoAgeAccel in predicting risk of overall cancer. Nevertheless, we also acknowledge several limitations. First, we calculated PhenoAge based on 9 biomarkers from blood, which were measured at baseline. As such, we were unable to access the change of PhenoAgeAccel during the follow-up period. Second, previous studies have indicated that patricians in the UK Biobank differ from the general UK population because of low participation and healthy volunteer bias (Fry et al., 2017). Finally, even though the findings were achieved from participants with diverse ethnic backgrounds of the UK Biobank, the generalizability of our findings should be further assessed in more diverse populations when available.

In summary, our study showed that accelerated aging, which was measured by PhenoAgeAccel, was consistently related to an increased risk of several site-specific cancer and overall cancer with adjustment for chronological age, within and across genetic risk groups. PhenoAgeAccel can serve as a productive tool to facilitate identification of cancer susceptible individuals, in combination with individual’s genetic background, and act as an intermediate phenotype to guide interventions for high risk groups and track intervention efficacy.

## Data Availability

All data produced in the present work are contained in the manuscript.

## Ethics approval and consent to participate

This research was conducted using the UK Biobank Resource (Application Number: 60169). UK Biobank has received ethics approval from the Research Ethics Committee (ref. 11/NW/0382).

## Availability of data and materials

The data underlying the results presented in the study are available from the UK Biobank (http://biobank.ndph.ox.ac.uk/showcase/).

## Authors’ Contributions

**L. Bian**: Data curation, formal analysis, validation, visualization, writing–original draft, writing–review and editing. **Z. Ma**: Data curation, formal analysis, validation, visualization, writing–review and editing. **X. Fu**: Data curation, validation, visualization, writing–review and editing. **C. Ji**: Data curation, validation, visualization. **T. Wang**: Methodology, formal analysis, validation, visualization, writing–review and editing. **C. Yan**: Methodology, writing–review and editing. **J. Dai**: Methodology, writing–review and editing. **H. Ma**: Supervision, project administration, writing–review and editing. **Z. Hu**: Conceptualization, supervision, project administration, writing–review and editing. **H. Shen**: Conceptualization, resources, supervision, project administration, writing–review and editing. **L. Wang**: Conceptualization, formal analysis, supervision, validation, writing–review and editing. **M. Zhu**: Conceptualization, formal analysis, supervision, funding acquisition, validation, writing–original draft, writing–review and editing. **G. Jin**: Conceptualization, formal analysis, supervision, funding acquisition, validation, writing–review and editing.

## Acknowledgments

The authors thank the investigators and participants in UK Biobank for their contributions to this study.

