## Appendix 1-table 1-11; Appendix 1-figure 1-2 for "Associations of Combined Phenotypic Aging and Genetic Risk with Incident Cancer: A Prospective Cohort Study"

**Additional Tables**

**Appendix 1-table 1. Association results of PhenoAgeAccel with site-specific cancer risk per 5 years increased**

|  |  |  | Model 1^a^ | |  | Model 2^b^ | |
| --- | --- | --- | --- | --- | --- | --- | --- |
| **Cancer site** | **N (cases)** | **Person-Years** | **HR (95% CI)** | ***P* value** |  | **HR (95% CI)** | ***P* value** |
| Lip, Oral Cavity, Pharynx | 374463 (445) | 2645366 | 1.19 (1.10-1.30) | 4.82×10^-5^ |  | 1.13 (1.04-1.24) | 4.52×10^-3^ |
| Oesophagus | 374463 (433) | 2645807 | 1.48 (1.37-1.59) | 2.27×10^-23^ |  | 1.41 (1.31-1.53) | 9.81×10^-18^ |
| Stomach | 374463 (292) | 2646114 | 1.39 (1.26-1.53) | 3.08×10^-11^ |  | 1.35 (1.22-1.49) | 2.90×10^-9^ |
| Colon, Rectum | 374463 (2623) | 2639167 | 1.19 (1.15-1.23) | 1.05×10^-21^ |  | 1.17 (1.13-1.21) | 9.89×10^-18^ |
| Pancreas | 374463 (518) | 2646135 | 1.30 (1.20-1.40) | 1.41×10^-11^ |  | 1.28 (1.18-1.38) | 4.42×10^-10^ |
| Lung, Bronchus, Trachea | 374463 (1754) | 2644034 | 1.48 (1.42-1.53) | 6.40×10^-94^ |  | 1.38 (1.33-1.44) | 2.04×10^-60^ |
| Skin Melanoma | 374463 (1145) | 2642902 | 0.98 (0.93-1.04) | 0.55 |  | 0.99 (0.93-1.05) | 0.72 |
| Breast | 201032 (4553) | 1409789 | 1.07 (1.04-1.10) | 1.52×10^-6^ |  | 1.06 (1.03-1.09) | 4.07×10^-5^ |
| Cervix Uteri | 201032 (59) | 1425046 | 1.27 (1.01-1.60) | 0.04 |  | 1.27 (1.00-1.60) | 0.05 |
| Corpus Uteri, NOS | 201032 (724) | 1422850 | 1.27 (1.19-1.36) | 2.99×10^-13^ |  | 1.24 (1.16-1.33) | 2.61×10^-10^ |
| Ovary | 201032 (476) | 1423963 | 1.10 (1.00-1.20) | 0.04 |  | 1.09 (1.00-1.19) | 0.06 |
| Prostate | 173431 (4736) | 1206048 | 0.94 (0.91-0.96) | 1.42×10^-5^ |  | 0.94 (0.92-0.97) | 1.82×10^-4^ |
| Testis | 173431 (58) | 1221139 | 1.14 (0.88-1.49) | 0.32 |  | 1.15 (0.88-1.51) | 0.3 |
| Kidney | 374463 (618) | 2645040 | 1.40 (1.31-1.49) | 1.78×10^-22^ |  | 1.36 (1.27-1.45) | 5.86×10^-18^ |
| Bladder | 374463 (524) | 2645064 | 1.29 (1.19-1.39) | 7.14×10^-11^ |  | 1.23 (1.13-1.33) | 2.75×10^-7^ |
| Brain, Central Nerves | 374463 (380) | 2646176 | 1.07 (0.97-1.18) | 0.18 |  | 1.06 (0.96-1.17) | 0.23 |
| Thyroid | 374463 (198) | 2645901 | 1.01 (0.88-1.16) | 0.90 |  | 1.01 (0.88-1.16) | 0.89 |
| Hodgkin’s Disease | 374463 (66) | 2646383 | 1.34 (1.08-1.64) | 6.54×10^-3^ |  | 1.30 (1.05-1.61) | 0.02 |
| Multiple Myeloma | 374463 (357) | 2645735 | 1.18 (1.07-1.29) | 8.67×10^-4^ |  | 1.17 (1.06-1.29) | 2.03×10^-3^ |
| Lymphoid Leukaemia | 374463 (323) | 2645604 | 1.28 (1.16-1.41) | 8.79×10^-7^ |  | 1.26 (1.14-1.39) | 6.51×10^-6^ |

^a^ Cox proportional hazards regression adjusted for age, height, cancer family history, Townsend deprivation index at recruitment, and the first 10 principal components of ancestry.

^b^ Cox proportional hazards regression adjusted for Model 1, as well as cancer polygenic risk score and healthy lifestyle.

Abbreviations: CI, confidence interval; HR, hazards ratio.

**Appendix 1-table 2. Sensitivity analysis of association between different risk levels of PhenoAgeAccel and cancer risk**

|  |  |  |  | Model 1^a^ | |  | Model 2^b^ | |
| --- | --- | --- | --- | --- | --- | --- | --- | --- |
|  |  | **N (cases)** | **Person-Years** | **HR (95% CI)** | ***P* value** |  | **HR (95% CI)** | ***P* value** |
| **Men** | **Tertiles** |  |  |  |  |  |  |  |
|  | Low (Q1) | 55498 (3132) | 386584 | Ref. |  |  | Ref. |  |
|  | Intermediate (Q2) | 58966 (3686) | 406761 | 1.07 (1.02-1.12) | 6.24×10^-3^ |  | 1.05 (1.00-1.10) | 0.03 |
|  | High (Q3) | 58967 (4714) | 397283 | 1.30 (1.24-1.36) | 4.16×10^-29^ |  | 1.26 (1.20-1.32) | 1.92×10^-22^ |
|  | ***P for trend*** |  |  | 1.07×10^-30^ |  |  | 8.82×10^-24^ |  |
|  | **Quartiles** |  |  |  |  |  |  |  |
|  | Low (Q1) | 43358 (2405) | 302638 | Ref. |  |  | R×10f. |  |
|  | Intermediate (Q2-Q3) | 86715 (5488) | 597359 | 1.10 (1.05-1.16) | 8.44×10^-5^ |  | 1.08 (1.03-1.14) | 1.38×10^-3^ |
|  | High (Q4) | 43358 (3639) | 290631 | 1.37 (1.30-1.44) | 1.93×10^-32^ |  | 1.32 (1.25-1.39) | 3.15×10^-25^ |
|  | ***P for trend*** |  |  | 1.56×10^-34^ |  |  | 5.21×10^-27^ |  |
| **Women** | **Tertiles** |  |  |  |  |  |  |  |
|  | Low (Q1) | 64330 (3099) | 450233 | Ref. |  |  | Ref. |  |
|  | Intermediate (Q2) | 68351 (3653) | 473668 | 1.16 (1.10-1.21) | 3.37×10^-9^ |  | 1.14 (1.09-1.20) | 1.01×10^-7^ |
|  | High (Q3) | 68351 (4086) | 469661 | 1.35 (1.29-1.42) | 8.26×10^-37^ |  | 1.31 (1.25-1.37) | 2.75×10^-28^ |
|  | ***P for trend*** |  |  | 4.32×10^-37^ |  |  | 1.85×10^-28^ |  |
|  | **Quartiles** |  |  |  |  |  |  |  |
|  | Low (Q1) | 50258 (2404) | 352128 | Ref. |  |  | Ref. |  |
|  | Intermediate (Q2-Q3) | 100516 (5312) | 696757 | 1.16 (1.10-1.22) | 1.83×10^-9^ |  | 1.14 (1.09-1.20) | 9.62×10^-8^ |
|  | High (Q4) | 50258 (3122) | 344677 | 1.44 (1.36-1.52) | 1.14×10^-40^ |  | 1.38 (1.31-1.46) | 1.15×10^-31^ |
|  | ***P for trend*** |  |  | 2.97×10^-41^ |  |  | 4.09×10^-32^ |  |

^a^ Cox proportional hazards regression adjusted for age, height, cancer family history, Townsend deprivation index at recruitment, and the first 10 principal components of ancestry.

^b^ Cox proportional hazards regression adjusted for Model 1, as well as cancer polygenic risk score and healthy lifestyle.

Abbreviations: CI, confidence interval; HR, hazards ratio; Ref, reference.

**Appendix 1-table 3. Sensitivity analysis of association between PhenoAgeAccel and cancer risk by excluding of patients diagnosed in the first two follow-up**

|  |  |  |  | Model 1^a^ | |  | Model 2^b^ | |
| --- | --- | --- | --- | --- | --- | --- | --- | --- |
|  |  | **N (cases)** | **Person-Years** | **HR (95% CI)** | ***P* value** |  | **HR (95% CI)** | ***P* value** |
| **Men** | **Per 5 years increase** | 170688 (8789) | 1187773 | 1.12 (1.10-1.15) | 1.73×10^-29^ |  | 1.11 (1.09-1.13) | 6.55×10^-23^ |
|  | **Category** |  |  |  |  |  |  |  |
|  | Biologically younger | 80205 (3612) | 563719 | Ref. |  |  | Ref. |  |
|  | Biologically older | 90483 (5177) | 624054 | 1.20 (1.15-1.25) | 2.07×10^-16^ |  | 1.17 (1.12-1.22) | 1.45×10^-12^ |
|  | **Quintiles** |  |  |  |  |  |  |  |
|  | Low (Q1) | 34138 (1464) | 240986 | Ref. |  |  | Ref. |  |
|  | Intermediate (Q2-Q4) | 102412 (5106) | 714091 | 1.12 (1.06-1.19) | 1.31×10^-4^ |  | 1.10 (1.04-1.17) | 1.42×10^-3^ |
|  | High (Q5) | 34138 (2219) | 232696 | 1.37 (1.28-1.46) | 4.23×10^-20^ |  | 1.31 (1.23-1.41) | 2.15×10^-15^ |
|  | ***P for trend*** |  |  | 2.57×10^-21^ |  |  | 2.21×10^-16^ |  |
| **Women** | **Per 5 years increase** | 198330 (8136) | 1390826 | 1.14 (1.12-1.16) | 4.65×10^-35^ |  | 1.12 (1.10-1.15) | 5.81×10^-27^ |
|  | **Category** |  |  |  |  |  |  |  |
|  | Biologically younger | 122113 (4694) | 860019 | Ref. |  |  | Ref. |  |
|  | Biologically older | 76217 (3442) | 530807 | 1.25 (1.20-1.31) | 1.72×10^-23^ |  | 1.22 (1.17-1.27) | 4.71×10^-18^ |
|  | **Quintiles** |  |  |  |  |  |  |  |
|  | Low (Q1) | 39666 (1492) | 280831 | Ref. |  |  | Ref. |  |
|  | Intermediate (Q2-Q4) | 118998 (4736) | 834654 | 1.11 (1.05-1.18) | 2.63×10^-4^ |  | 1.09 (1.03-1.16) | 2.48×10^-3^ |
|  | High (Q5) | 39666 (1908) | 275341 | 1.43 (1.34-1.53) | 7.52×10^-25^ |  | 1.37 (1.28-1.47) | 5.30×10^-19^ |
|  | ***P for trend*** |  |  | 2.14×10^-25^ |  |  | 1.80×10^-19^ |  |

^a^ Cox proportional hazards regression adjusted for age, height, cancer family history, Townsend deprivation index at recruitment, and the first 10 principal components of ancestry.

^b^ Cox proportional hazards regression adjusted for Model 1, as well as cancer polygenic risk score and healthy lifestyle.

Abbreviations: CI, confidence interval; HR, hazards ratio; Ref, reference.

**Appendix 1-table 4. Sensitivity analysis of association between PhenoAgeAccel and cancer risk in unimputed data**

|  |  |  |  | Model 1^a^ | |  | Model 2^b^ | |
| --- | --- | --- | --- | --- | --- | --- | --- | --- |
|  |  | **N (cases)** | **Person-Years** | **HR (95% CI)** | ***P* value** |  | **HR (95% CI)** | ***P* value** |
| **Men** | **Per 5 years increase** | 91265 (5569) | 628642 | 1.14 (1.11-1.17) | 5.33×10^-21^ |  | 1.13 (1.10-1.16) | 1.20×10^-17^ |
|  | **Category** |  |  |  |  |  |  |  |
|  | Biologically younger | 46495 (2518) | 323569 | Ref. |  |  | Ref. |  |
|  | Biologically older | 44770 (3051) | 305073 | 1.19 (1.13-1.25) | 1.24×10^-10^ |  | 1.17 (1.11-1.23) | 8.46×10^-9^ |
|  | **Quintiles** |  |  |  |  |  |  |  |
|  | Low (Q1) | 18253 (935) | 127782 | Ref. |  |  | Ref. |  |
|  | Intermediate (Q2-Q4) | 54759 (3216) | 377865 | 1.11 (1.03-1.19) | 4.94×10^-3^ |  | 1.09 (1.02-1.18) | 1.54×10^-2^ |
|  | High (Q5) | 18253 (1418) | 122994 | 1.37 (1.26-1.49) | 9.03×10^-14^ |  | 1.33 (1.23-1.45) | 1.49×10^-11^ |
|  | ***P for trend*** |  |  | 1.03×10^-14^ |  |  | 2.08×10^-12^ |  |
| **Women** | **Per 5 years increase** | 90400 (4517) | 626559 | 1.16 (1.12-1.19) | 1.80×10^-21^ |  | 1.14 (1.11-1.18) | 2.45×10^-18^ |
|  | **Category** |  |  |  |  |  |  |  |
|  | Biologically younger | 59095 (2777) | 411414 | Ref. |  |  | Ref. |  |
|  | Biologically older | 31305 (1740) | 215145 | 1.28 (1.21-1.36) | 4.29×10^-16^ |  | 1.26 (1.19-1.34) | 6.88×10^-14^ |
|  | **Quintiles** |  |  |  |  |  |  |  |
|  | Low (Q1) | 18080 (817) | 126772 | Ref. |  |  | Ref. |  |
|  | Intermediate (Q2-Q4) | 54240 (2676) | 375725 | 1.16 (1.07-1.26) | 1.81×10^-4^ |  | 1.15 (1.06-1.24) | 6.13×10^-4^ |
|  | High (Q5) | 18080 (1024) | 124062 | 1.44 (1.31-1.58) | 1.40×10^-14^ |  | 1.39 (1.27-1.53) | 2.61×10^-12^ |
|  | ***P for trend*** |  |  | 9.88×10^-15^ |  |  | 1.93×10^-12^ |  |

^a^ Cox proportional hazards regression adjusted for age, height, cancer family history, Townsend deprivation index at recruitment, and the first 10 principal components of ancestry.

^b^ Cox proportional hazards regression adjusted for Model 1, as well as cancer polygenic risk score and healthy lifestyle.

Abbreviations: CI, confidence interval; HR, hazards ratio; Ref, reference.

**Appendix 1-table 5. Sensitivity analysis of association between PhenoAgeAccel and cancer risk in the unrelated white British population**

|  |  |  |  | Model 1^a^ | |  | Model 2^b^ | |
| --- | --- | --- | --- | --- | --- | --- | --- | --- |
|  |  | **N (cases)** | **Person-Years** | **HR (95% CI)** | ***P* value** |  | **HR (95% CI)** | ***P* value** |
| **Men** | **Per 5 years increase** | 105888 (7193) | 726811 | 1.14 (1.11-1.16) | 7.91×10^-29^ |  | 1.12 (1.10-1.15) | 4.71×10^-23^ |
|  | **Category** |  |  |  |  |  |  |  |
|  | Biologically younger | 49976 (2954) | 347479 | Ref. |  |  | Ref. |  |
|  | Biologically older | 55912 (4239) | 379332 | 1.20 (1.15-1.26) | 1.99×10^-14^ |  | 1.18 (1.12-1.23) | 2.93×10^-11^ |
|  | **Quintiles** |  |  |  |  |  |  |  |
|  | Low (Q1) | 21178 (1182) | 148133 | Ref. |  |  | Ref. |  |
|  | Intermediate (Q2-Q4) | 63532 (4163) | 437212 | 1.14 (1.07-1.21) | 1.01×10^-4^ |  | 1.12 (1.05-1.19) | 9.70×10^-4^ |
|  | High (Q5) | 21178 (1848) | 141466 | 1.40 (1.30-1.51) | 2.76×10^-19^ |  | 1.35 (1.25-1.45) | 3.39×10^-15^ |
|  | ***P for trend*** |  |  | 1.80×10^-20^ |  |  | 3.48×10^-16^ |  |
| **Women** | **Per 5 years increase** | 118597 (6464) | 822885 | 1.15 (1.12-1.17) | 1.25×10^-30^ |  | 1.13 (1.10-1.16) | 6.69×10^-24^ |
|  | **Category** |  |  |  |  |  |  |  |
|  | Biologically younger | 73575 (3743) | 513046 | Ref. |  |  | Ref. |  |
|  | Biologically older | 45022 (2721) | 309839 | 1.26 (1.20-1.32) | 1.09×10^-19^ |  | 1.22 (1.16-1.29) | 2.89×10^-15^ |
|  | **Quintiles** |  |  |  |  |  |  |  |
|  | Low (Q1) | 23720 (1162) | 166407 | Ref. |  |  | Ref. |  |
|  | Intermediate (Q2-Q4) | 71157 (3779) | 493911 | 1.14 (1.07-1.22) | 1.00×10^-4^ |  | 1.12 (1.05-1.20) | 7.89×10^-4^ |
|  | High (Q5) | 23720 (1523) | 162567 | 1.45 (1.34-1.57) | 2.45×10^-21^ |  | 1.39 (1.28-1.50) | 1.33×10^-16^ |
|  | ***P for trend*** |  |  | 7.87×10^-22^ |  |  | 5.34×10^-17^ |  |

^a^ Cox proportional hazards regression adjusted for age, height, cancer family history, Townsend deprivation index at recruitment, and the first 10 principal components of ancestry.

^b^ Cox proportional hazards regression adjusted for Model 1, as well as cancer polygenic risk score and healthy lifestyle.

Abbreviations: CI, confidence interval; HR, hazards ratio; Ref, reference.

**Appendix 1-table 6. Sensitivity analysis of association between PhenoAgeAccel and cancer risk using retrained** **PhenoAge in cancer-free participants**

|  |  |  |  | **Model 1^a^** | |  | **Model 2^b^** | |
| --- | --- | --- | --- | --- | --- | --- | --- | --- |
|  |  | **N (cases)** | **Person-Years** | **HR (95% CI)** | ***P* value** |  | **HR (95% CI)** | ***P* value** |
| **Men** | **Per 5 years increase** | 173431 (11532) | 1190628 | 1.13 (1.11-1.15) | 8.56×10^-40^ |  | 1.11(1.09-1.14) | 1.62×10^-30^ |
|  | **Category** |  |  |  |  |  |  |  |
|  | Biologically younger | 78296(4649) | 544237 | Ref. |  |  | Ref. |  |
|  | Biologically older | 95135(6883) | 646391 | 1.13(1.09-1.17) | 4.76×10^-10^ |  | 1.10(1.06-1.14) | 1.57×10^-6^ |
|  | **Quintiles^c^** |  |  |  |  |  |  |  |
|  | Low | 34687 (1914) | 242497 | Ref. |  |  | Ref. |  |
|  | Intermediate | 104058 (6825) | 716012 | 1.11(1.06-1.17) | 2.94×10^-5^ |  | 1.09(1.04-1.15) | 6.84×10^-4^ |
|  | High | 34686 (2793) | 232119 | 1.29(1.22-1.37) | 2.56×10^-17^ |  | 1.24(1.16-1.31) | 3.71×10^-12^ |
|  | ***P for trend*** |  |  | 5.54×10^-18^ |  |  | 1.25×10^-12^ |  |
| **Women** | **Per 5 years increase** | 201032 (10838) | 1393562 | 1.16(1.14-1.18) | 9.08×10^-54^ |  | 1.14(1.12-1.16) | 3.15×10^-41^ |
|  | **Category** |  |  |  |  |  |  |  |
|  | Biologically younger | 121477(6229) | 846419 | Ref. |  |  | Ref. |  |
|  | Biologically older | 79555(4609) | 547143 | 1.20(1.15-1.24) | 4.20×10^-20^ |  | 1.16(1.12-1.21) | 8.13×10^-14^ |
|  | **Quintiles^c^** |  |  |  |  |  |  |  |
|  | Low | 40207 (2005) | 281830 | Ref. |  |  | Ref. |  |
|  | Intermediate | 120618 (6303) | 836609 | 1.10(1.05-1.16) | 1.69×10^-4^ |  | 1.08(1.03-1.14) | 2.78×10^-3^ |
|  | High | 40207 (2530) | 275123 | 1.40(1.32-1.48) | 6.36×10^-29^ |  | 1.33(1.25-1.41) | 6.59×10^-21^ |
|  | ***P for trend*** |  |  | 1.48×10^-29^ |  |  | 2.01×10^-21^ |  |

^a^ Cox proportional hazards regression adjusted for age, height, cancer family history, Townsend deprivation index at recruitment, and the first 10 principal components of ancestry.

^b^ Cox proportional hazards regression adjusted for Model 1, as well as cancer polygenic risk score and healthy lifestyle.

^c^ Defined by quintiles of PhenoAgeAccel: low (the bottom quintile), intermediate (quintiles 2-4) and high (the top quintile).

Abbreviations: CI, confidence interval; HR, hazards ratio; Ref, reference.

**Appendix 1-table 7. RERI and AP for additive interaction between genetic and PhenoAgeAccel categories^a^**

| **Genetic risk^b^** | | **Biologically older** | |
| --- | --- | --- | --- |
|  |  | **RERI (95%CI)** | **AP (95%CI)** |
| **Men** | Intermediate | 0.048 (-0.088,0.167) | 0.029 (-0.052,0.106) |
|  | High | -0.112 (-0.315,0.078) | -0.049 (-0.135,0.033) |
| **Women** | Intermediate | 0.095 (-0.036,0.213) | 0.062 (-0.023,0.142) |
|  | High | 0.053 (-0.127,0.229) | 0.027 (-0.067,0.116) |

^a^ Adjusted for age, height, cancer family history, Townsend deprivation index at recruitment, and the first 10 principal components of ancestry.

^b^ Participants were divided into low (the bottom quintile of CPRS), intermediate (quintile 2 to 4) or high (the top quintile) genetic risk.

Abbreviations: RERI, relative excess risk due to interaction; AP, attributable proportion due to interaction; CI, confidence interval.

**Appendix 1-table 8. Risk of incident cancer according to PhenoAgeAccel categories within each genetic risk level ^a^**

| **Gender** | **PhenoAgeAccel**  **category^b^** | **Low genetic risk** | | |  | **Intermediate genetic risk** | | |  |  | **High genetic risk** |  |
| --- | --- | --- | --- | --- | --- | --- | --- | --- | --- | --- | --- | --- |
|  |  | **Low** | **Intermediate** | **High** |  | **Low** | **Intermediate** | **High** |  | **Low** | **Intermediate** | **High** |
| Men | No. of cases/Person years | 256/50601 | 946/142730 | 497/46158 |  | 1041/144491 | 3845/431551 | 1732/139690 |  | 567/47427 | 1877/142032 | 771/45949 |
|  | Hazards ratio  (95% CI) | Ref. | 1.22  (1.06-1.40) | 1.78  (1.53-2.07) |  | Ref. | 1.18  (1.10-1.26) | 1.48  (1.37-1.60) |  | Ref. | 1.04  (0.94-1.14) | 1.21  (1.09-1.36) |
|  | *P* value |  | 4.96×10^-3^ | 1.23×10^-13^ |  |  | 2.43×10^-6^ | 3.49×10^-23^ |  |  | 0.44 | 4.92×10^-4^ |
|  | Absolute risk (%)-  5 years (95% CI) | 2.36  (2.06-2.66) | 3.09  (2.87-3.32) | 5.04  (4.57-5.51) |  | 3.34  (3.13-3.55) | 4.14  (3.99-4.29) | 5.77  (5.49-6.06) |  | 5.56  (5.09-6.04) | 6.16  (5.85-6.48) | 7.85  (7.26-8.43) |
|  | Absolute risk increase (%)-  5 years (95% CI) | Ref. | 0.73  (0.37-1.09) | 2.68  (2.13-3.22) |  | Ref. | 0.80  (0.55-1.03) | 2.44  (2.07-2.78) |  | Ref. | 0.60  (0.04-1.16) | 2.28  (1.57-2.97) |
| Women | No. of cases/Person years | 327/56764 | 1033/168417 | 431/54950 |  | 1127/170088 | 3691/502281 | 1465/164994 |  | 492/55060 | 1627/165643 | 645/55365 |
|  | Hazards ratio  (95% CI) | Ref. | 1.11  (0.98-1.26) | 1.48  (1.28-1.71) |  | Ref. | 1.16  (1.08-1.24) | 1.47  (1.36-1.59) |  | Ref. | 1.14  (1.03-1.26) | 1.44  (1.28-1.62) |
|  | *P* value |  | 0.09 | 1.10×10^-7^ |  |  | 2.26×10^-5^ | 6.14×10^-22^ |  |  | 0.01 | 1.65×10^-9^ |
|  | Absolute risk (%)-  5 years (95% CI) | 2.74  (2.43-3.05) | 2.92  (2.72-3.12) | 3.73  (3.36-4.11) |  | 3.14  (2.94-3.33) | 3.48  (3.36-3.61) | 4.21  (3.99-4.44) |  | 4.29  (3.89-4.68) | 4.71  (4.45-4.97) | 5.60  (5.14-6.05) |
|  | Absolute risk increase (%)-  5 years (95% CI) | Ref. | 0.18  (-0.17-0.53) | 1.00  (0.50-1.44) |  | Ref. | 0.35  (0.04-1.16) | 1.08  (0.80-1.38) |  | Ref. | 0.43  (0.00-0.91) | 1.31  (0.73-1.89) |

^a^ Cox proportional hazards regression is adjusted for age, height, family history of cancer, Townsed deprivation index, height and the first 10 principal components of ancestry.

^b^ Defined by quintiles of PhenoAgeAccel: low (the bottom quartile), intermediate (quintiles 2-4) and high (the top quintile).

Abbreviations: CI, confidence interval; Ref, reference.

**Appendix 1-table 9. PhenoAgeAccel of participants stratified by lifestyle factors**

|  | **PhenoAgeAccel in men,**  **median (IQR), years** |  | **PhenoAgeAccel in women,**  **median (IQR), years** |
| --- | --- | --- | --- |
| **Smoking status** |  |  |  |
| Smoker | 0.98 (-1.96-4.39) |  | -1.03 (-4.00-2.42) |
| Non smoker | -0.17 (-2.79-2.76) |  | -1.58 (-4.41-1.67) |
| **Alcohol drinking status** |  |  |  |
| Drinker | 0.32 (-2.43-3.51) |  | -1.43 (-4.30-1.87) |
| Non drinker | 0.94 (-2.20-4.50) |  | -0.40 (-3.64-3.47) |
| **Body mass index** |  |  |  |
| Unfavorable | 1.82 (-1.07-5.30) |  | 0.96 (-2.11-4.58) |
| Favorable | -0.14 (-2.80-2.92) |  | -2.04 (-4.76-1.02) |
| **Physical activity** |  |  |  |
| Unfavorable | 0.70 (-2.14-4.11) |  | -0.98 (-3.96-2.54) |
| Favorable | 0.16 (-2.56-3.23) |  | -1.59 (-4.41-1.63) |
| **Diet** |  |  |  |
| Unfavorable | 0.54 (-2.22-3.74) |  | -1.11 (-4.00-2.24) |
| Favorable | -0.48 (-3.16-2.68) |  | -2.00 (-4.82-1.24) |
| **Lifestyle category ^a^** |  |  |  |
| Unfavorable | 1.78 (-1.21-5.37) |  | 0.37 (-2.73-4.03) |
| Intermediate | -0.01 (-2.67-3.02) |  | -1.62 (-4.43-1.56) |
| Favorable | -0.97 (-3.53-1.81) |  | -2.50 (-5.15-0.56) |

^a^ Participants were divided into favorable (at least 4 healthy lifestyle factors), intermediate (2 or 3 healthy lifestyle factors), or unfavorable (0 or 1 healthy lifestyle factor) according to the number of healthy lifestyle factors.

**Appendix 1-table 10. Association results of PhenoAgeAccel with lifestyle factors of participants^a^**

|  | **PhenoAgeAccel in men** | |  | **PhenoAgeAccel in women** | |
| --- | --- | --- | --- | --- | --- |
|  | **Beta coefficients** | ***P* value** |  | **Beta coefficients** | ***P* value** |
| **Healthy lifestyle factors** |  |  |  |  |  |
| No current smoking | -1.10 | <0.001 |  | -0.57 | <0.001 |
| No alcohol consumption | 0.58 | <0.001 |  | 1.01 | <0.001 |
| Normal BMI | -2.09 | <0.001 |  | -3.11 | <0.001 |
| Regular physical activity | -0.74 | <0.001 |  | -0.68 | <0.001 |
| Healthy diet | -1.07 | <0.001 |  | -0.90 | <0.001 |
| **Number of healthy**  **lifestyle factors** | -1.01 | <0.001 |  | -0.98 | <0.001 |

^a^ Adjusted for age, height, family cancer history, Townsend deprivation index and the first 10 principal components of ancestry.

**Appendix 1-table 11. RERI and AP for additive interaction between genetic and lifestyle factor on PhenoAgeAccel ^a^**

| **Lifestyle category^b^** | | **Genetic risk ^c^** | | | | |
| --- | --- | --- | --- | --- | --- | --- |
|  |  | **Intermediate** | |  | **High** | |
|  |  | **RERI (95%CI)** | **AP (95%CI)** |  | **RERI (95%CI)** | **AP (95%CI)** |
| **Men** | Intermediate | 0.07(-0.02,0.15) | 0.04(-0.01,0.10) |  | 0.02(-0.11,0.13) | 0.01(-0.06,0.07) |
|  | Unfavorable | 0.06(-0.04,0.16) | 0.03(-0.02,0.08) |  | -0.04(-0.18,0.10) | -0.02(-0.08,0.04) |
| **Women** | Intermediate | 0.02(-0.07,0.10) | 0.01(-0.04,0.06) |  | 0.04(-0.07,0.15) | 0.02(-0.03,0.07) |
|  | Unfavorable | 0.09(-0.02,0.20) | 0.04(-0.01,0.08) |  | 0.06(-0.09,0.21) | 0.02(-0.03,0.07) |

^a^ Adjusted for age, height, cancer family history, Townsend deprivation index at recruitment, and the first 10 principal components of ancestry.

^b^ Participants were divided into favorable (at least 4 healthy lifestyle factors), intermediate (2 or 3 healthy lifestyle factors), or unfavorable (0 or 1 healthy lifestyle factor) according to the number of healthy lifestyle factors.

^c^ Participants were divided into low (the bottom quintile of PhenoAgeAccel PRS), intermediate (quintile 2 to 4) or high (the top quintile) genetic risk.

Abbreviations: RERI, relative excess risk due to interaction; AP, attributable proportion due to interaction; CI, confidence interval.

**Additional Figures**

**Appendix 1-figure 1. Flowchart for filtering participants from the UK Biobank cohort**

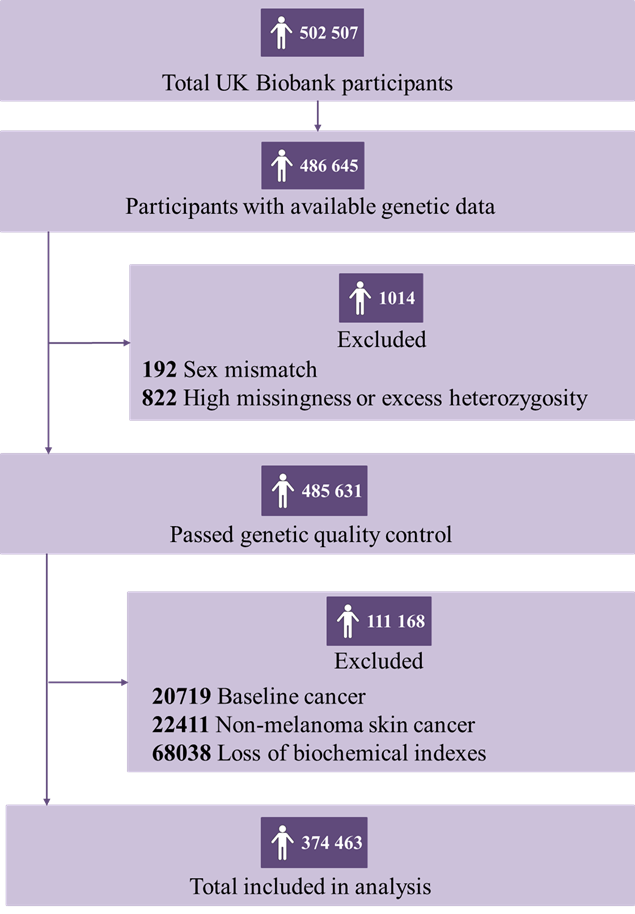

**Appendix 1-figure 2. Risk of incident cancer according to genetic and PhenoAgeAccel categories in the UKB cohort for (top) men and (bottom) women.** The HRs were estimated using Cox proportional hazard models with adjustment for age, height, family history of cancer, Townsend deprivation index, and the first 10 principal components of ancestry. Participants were divided into low (the bottom quintile of PhenoAgeAccel), intermediate (quintiles 2-4), and high (the top quintile) accelerated aging under different genetic risk groups.

**Men**

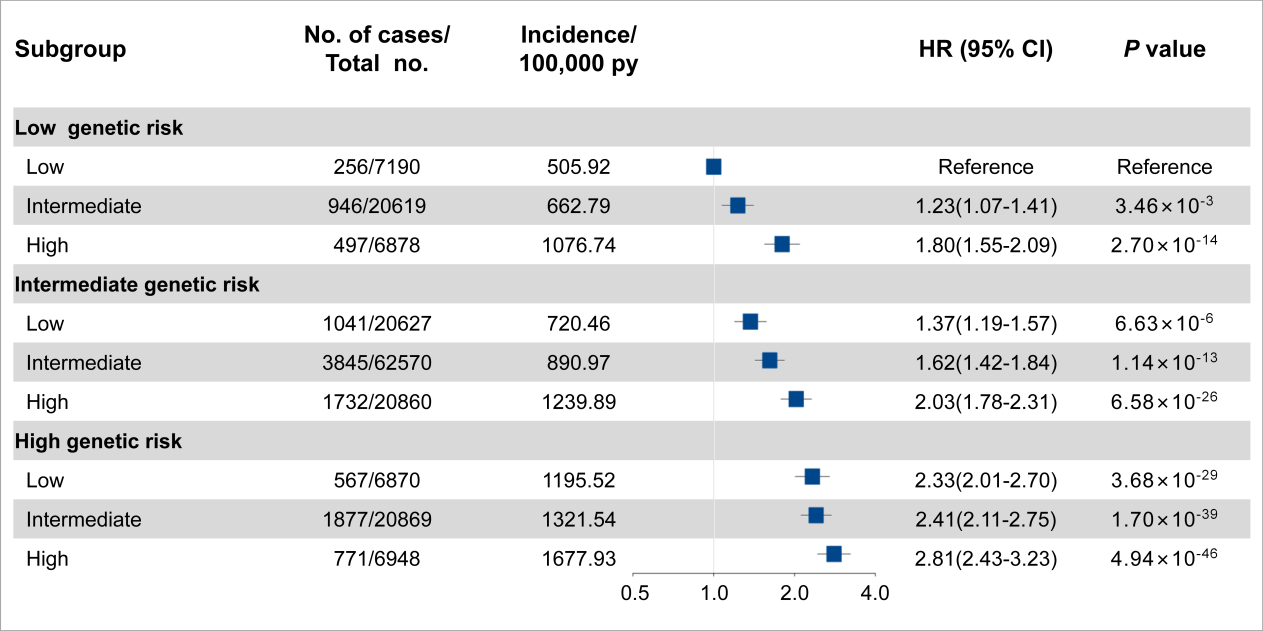

**Women**

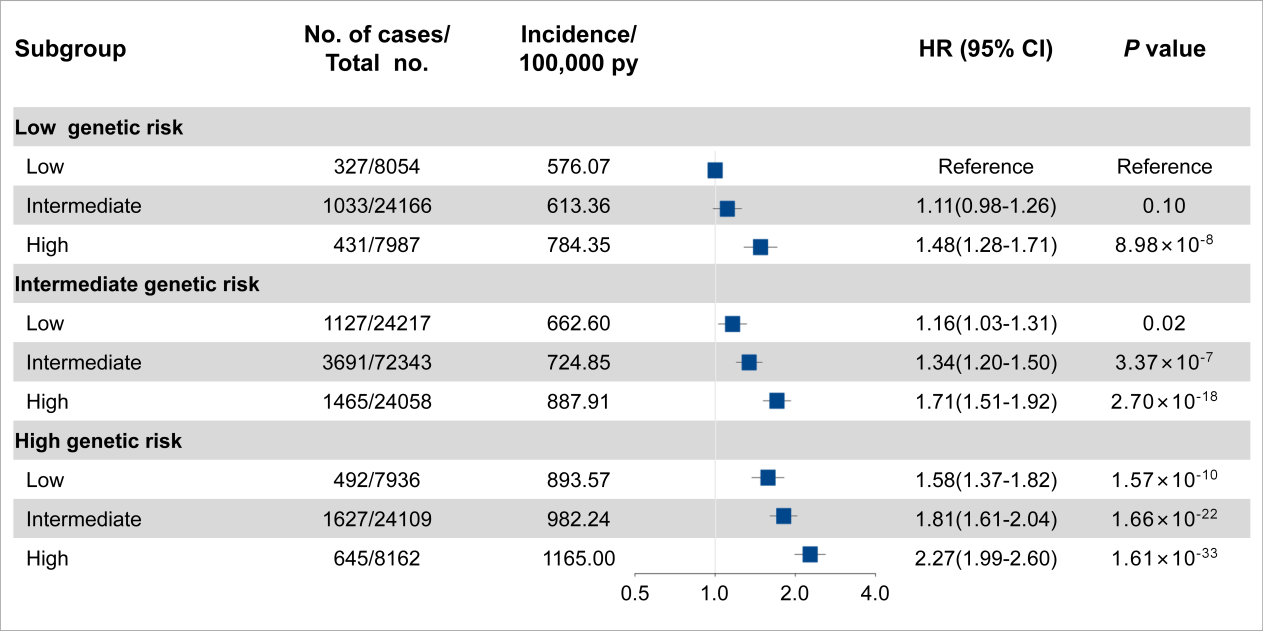
